# Signature transcriptome analysis of stage specific atherosclerotic plaques of patients

**DOI:** 10.1101/2021.06.15.21259006

**Authors:** Sonia Verma, Abhay Kumar, Rajiv Narang, Akshay K Bisoi, Dipendra K Mitra

## Abstract

**Background:** Inflammation plays an important role in all the stages of atherosclerotic plaque development. The current study aimed at assessing the altered expression of genes functioning in inflammation within the early stage (ES)and advanced stage (AS) atherosclerotic plaques obtained from patients undergoing coronary artery bypass grafting (CABG) surgery and identifying biomarker panel/sthat may detect the status of plaque stages using peripheral blood samples.

**Methods:** A section of ES and ASplaques and normal left internal mammary arteries (LIMA) were obtained from 8 patients undergoing theCABG surgery. Total RNA isolated was analysed for mRNA and miRNAexpression profile by Affymetrix arrays. Significant number of mRNAs was found to be differentially expressed in ES and ASplaque tissues relative to LIMA. The pathway analysis of differentially expressed mRNAs in the two plaque stages was also performed using DAVID Bioinformatics Database.

**Results:** The mRNAs were found to be involved in critical inflammatory processes such as Toll-like receptor signalling pathway and cytokine-cytokine receptor interaction. Few miRNAs targeting these mRNAs were also altered in the two plaque conditions. QRT-PCR results showedsimilar expression pattern of few of themRNAs and miRNAsin peripheral blood of same patients relative to healthy controls.

**Conclusion:** Changes in mRNA and miRNA expression associated with various inflammatory processes occur in different atherosclerotic stage plaques as well as peripheral blood. Detection of such variations in patients’ blood can be used as a possible prognostic tool to detect and/or predict the risk and stage of atherosclerosis.

## Introduction

Atherosclerotic vascular plaques are major pathological basis for cardiovascular diseases (CVD) including ischemic heart disease (IHD), stroke, peripheral vascular disease and renovascular hypertension. Various effector immune mechanisms and signalling networks are involved in the pathology of atherosclerotic plaque formation[1–4]. The process starts with inflammatory activation of endothelial cells followed by migration of effector immune cells like monocytes and leukocytes into the atheroma under the influence of various homing molecules [5]. In the inflammatory milieu of atheroma, monocytes transform into macrophages and phagocytise lipoprotein particles to generate foamy cells, a hallmark of nascent atheroma[6]. In evolving atheroma, abundance of secretory inflammatory mediators such as cytokines, reactive oxygen species, perpetuate the local inflammatory response [7]. Inflammation driven plaque formation triggers a battery of subsequent complications including rupture of the thrombotic plaque and/or blockade of vascular lumen leading to ischemic pathologies [8].

As the vascular plaque evolves, it converts from an early, lipid-rich atheroma to advanced, fibro-calcified lesion[9, 10]. Lipid-rich plaques with a thin fibrous cap are prone to rupture, precipitating in thrombus formation, leading to vascular occlusion causing acute ischemia and infarction of tissue. Methods to determine nature of plaque are generally invasive and cumbersome. However, despite major advances in treatment, significant numbers of coronary artery disease (CAD) patients develop acute coronary syndromes and even have sudden cardiac death. Early detection of vulnerable plaque can identify patients at high risk for such events. Therefore it has become critical to develop improved molecular signature profiles for screening the plaque formation, preferably with stage-specific signatures. Development of improved prognostics biomarker(s) and thus, novel therapeutic strategies for CVD, necessitates better understanding and identification of various key molecules and genetic pathways uniquely involved in various stages of atherosclerotic plaque formation.

Attempts towards analysis of the gene expression profiles in atherosclerotic plaques are reported [11–17]. For comparative study, these reports used samples from different individuals, which may confound the results due to inter-individual genetic variation, tissue-specific differences in transcriptomes, and differences in systemic parameters. In CAD patients, distinct regions with ES or AS plaques may coexist within a single artery, hinting towards pathogenic attribute(s) leading to their sequential and/or differential evolution[18, 19]. Previous studies are mostly based on using autopsied specimen which may not truly represent the in vivo changes of the plaque. Therefore, to have better insights in to the molecular profile of gene expression in the various stages atherosclerotic plaques, we performed microarray analysis of early stage (ES) and advanced stage (AS) plaques as well as healthy Left Internal Mammary Artery (LIMA) tissues obtained surgically from the same live individuals undergoing coronary artery bypass grafting (CABG) surgery.

We identified several inflammatory processes and pro-inflammatory cytokines associated with these plaques, evidencing a sustained inflammation and cytokine storm in the patients. Also, the miRNA targeting these genes were found to be differentially expressed in the respective plaque stage. The expression levels of some of these genes were found to be following the same trend in patients’ blood as observed in the plaques. Conclusively, our data provides for an early and faster prognostic tool for detecting and predicting the risk and stage of atherosclerosis.

## Material and Methods

### Tissue Collection

Patients’ samples were obtained from 8 individuals (males-5, females-3), aged 55-80 years, undergoing CABG surgery at the apex tertiary referral centre, All India Institute of Medical Sciences (AIIMS), New Delhi, India.Plaque tissues excised at the time of surgery as well as a 5-8 mm segment of LIMA was taken for analysis. The plaque tissue was divided into two parts depending upon extent of calcification and lipid content. We used ES/lipid-rich plaque and AS/fibro-calcified plaque for further study. All these tissue fractions and LIMA segment were frozen immediately in QIAzol and stored at −80°C before RNA extraction. Written informed consent was obtained from each patient included in the study. The study protocol conforms to the ethical guidelines of the 1975 Declaration of Helsinki and has been priorly approved by the Institution’s ethics committee on research on humans.

### RNA Isolation

RNA was extracted from tissues using QiagenmiRNeasy Mini Kit. Briefly, the tissue samples were homogenized in QIAzolLysis Reagent. The homogenates were separated into aqueous and organic phases by centrifugation after adding chloroform. The upper, aqueous phases containing RNA were extracted. After adding ethanol, the samples were applied to the RNeasy Mini spin column for total RNA binding to the membrane. Following washes, total RNAs were eluted in RNase-free water. The concentrations of the RNA were determined using NanoDrop 2000 (Thermo Scientific, USA). RNA from peripheral blood was also isolated using the same kit.

### Microarray Analysis

For microarray analysis, RNA from tissues with similar condition of 4 patients was pooled to make one batchfor sequencing. Like this, two batches, each having 3 different samples, representing ES, AS and LIMA, were sent for sequencing to Imperialls Life Sciences (P) limited, India. In brief, for mRNA, cDNA synthesis, amplification, and gene expression profiling were done with the GeneChip WT PLUS Reagent Kit (Affymetrix) and labelled with the Genechip hybridization wash and stain kit (Affymetrix). Samples were hybridized with Gene Chip Human Transcriptome Array (HTA) 2.0 (Affymetrix) following manufacturer’s protocol. For miRNA, total RNA was labelled with Biotin using the AffymetrixFlashTag™ Biotin HSR kit following the manufacturer’s protocol. Labelled extracts were hybridized to the GeneChipmiRNA 4.0 Arrays overnight and subsequently processed usingGeneChip Hybridization Wash and Stain kit (Affymetrix) according to manufacturer’s instructions. Data analysis was done using appropriate software.

### Functional Enrichment of genes

The online tool, DAVID Bioinformatics Resources analyses was used to conduct the Kyoto Encyclopedia of Genes and Genomes (KEGG) pathway enrichment analysis of statistically significant, differentially-expressed mRNAs. The *p* < 0.05 was set as the cut-off for selecting significantly over-represented pathways.

### miRNAs targeting differentially expressed mRNAs

Differentially expressed mRNAs were subjected to the online database miRWalk(http://mirwalk.uni-hd.de/),where correlations between miRNAs and their target genes have been experimentally confirmed. In the database, three algorithms including targetScan, miRDB and miRTarBase are used to predict miRNAs targeting gene of interest. Any miRNA predicted for a given mRNA by all the three algorithms, was considered to be targeting that gene.

### miRNA-target genes correlation

The reverse correlation between mRNAs and the miRNAs targeting these genes was screened by overlapping the predicted miRNAs of up-regulated mRNAs with the identified down-regulated miRNAs and vice-versa for the respective plaque tissue relative to LIMA.

### QRT-PCR

1μg of RNA isolated from peripheral blood was reverse transcribed and used for miRNA quantification using Mir-X™ miRNA First-Strand Synthesis and TB Green qRT-PCR kit (Takara Bio Inc.). U6 was used as an internal control. For mRNA quantification, about 1μg of RNA isolated from peripheral blood was converted to cDNA with the cDNA Reverse Transcription Kit (Applied Biosystems, USA). Quantitative RT-PCR (qRT-PCR) was carried out on RealPlex PCR system (Eppendorf, USA) with SYBR Premix Ex Taq™ (TaKaRa Bio Inc.). GAPDH was used as internal control. Statistical analysis was performed using GraphPad 7.0. The primers used are enlisted in **Supplementary Table 1**.

### Data Availability

Raw data are available from the European Bioinformatics Institute ArrayExpress data repository (accession number-E-MTAB-10052 (mRNA data); E-MTAB-10051 (miRNA data)).

## Results

### Differential gene expression profiling in ES and AS plaque

To investigate the variation in the expression of genes in human ES and AS atherosclerotic plaques, microarray sequencing and analysis of total RNA isolated from both the plaque tissues as well as LIMA was carried out. For ES plaques, 518 genes were significantly up-regulated and 497 genes were significantly down-regulated relative to LIMA **(Figure 1A)**. In AS plaque samples, we found more number of genes to be differentially expressed, i.e. 846 up-regulated genes while 1338 down-regulated genes as compared to LIMA **(Figure 1A)**. Interestingly, in both the plaques 279 and 358 common genes were up and down-regulated, respectively**(Figure 1A)**. Some of these genes such asSPP1, TGFB, and AKT3 have been associated with atherosclerosis in previous studies on mice and human samples, thus supporting our findings[20–25]. Topmost 10 up-regulated and down-regulated mRNAs are presented in **Figure 1B and 1C**.In summary, the microarray data indicates that atherosclerotic plaques are associated with alterations in gene expression which may further differ amongvarious plaque stages.

**FIGURE 1:**
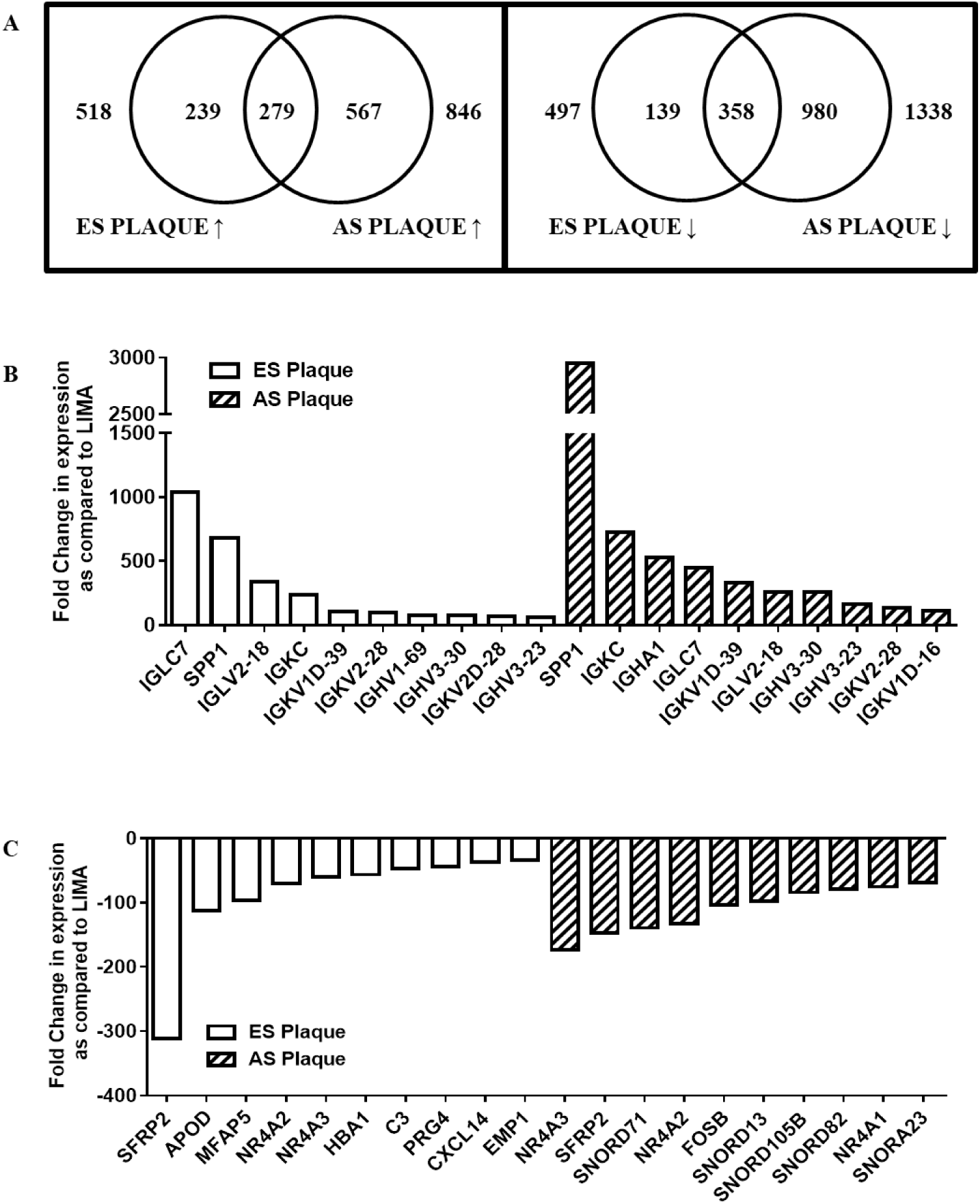
Microarray analysis revealsalterations in gene expression associated with atherosclerotic plaques. (**A)** Venn diagram of differentially expressed genes in early (ES) and advanced stage (AS) atherosclerotic plaqueas compared to Left Internal Mammary Artery (LIMA) (↑-up-regulated; ↓-down-regulated).**(B)** Bar graph of top 10 mRNAs significantly up-regulated as compared to LIMA in microarray analysis. **(C)**Bar graph of top 10 mRNAs significantly down-regulated as compared to LIMA in microarray analysis. *n=2(each with sample pooled from 4 patients); p<0*.*05*

### ES Plaques are associated with up-regulation of inflammatory mediators

Inflammation plays an important role in atherosclerosis. Elevated levels of inflammatory cytokines and chemokines have been observed in various studies on affected cardiac tissues [11, 12, 14– 17]. In the current study, we delineated the genes involved in the inflammation of ES and AS plaques by KEGG pathway analysis of differentially expressed genes using DAVID. In ES plaques, 19up-regulated genes were related to chemokine signalling pathway and cytokine-cytokine receptor interaction**(Figure 2A, Supplementary Table 2)**. Interestingly, most of these pro-inflammatory genes like CCL5, CXCL10, CXCL9, CTSK and CD14 have been previously reported to be elevated in the serum of patients with coronary artery diseases[11, 12, 14–17]. The list of differentially expressed genes functioning in inflammation pathways is shown in **Figure 2C**.

**FIGURE 2:**
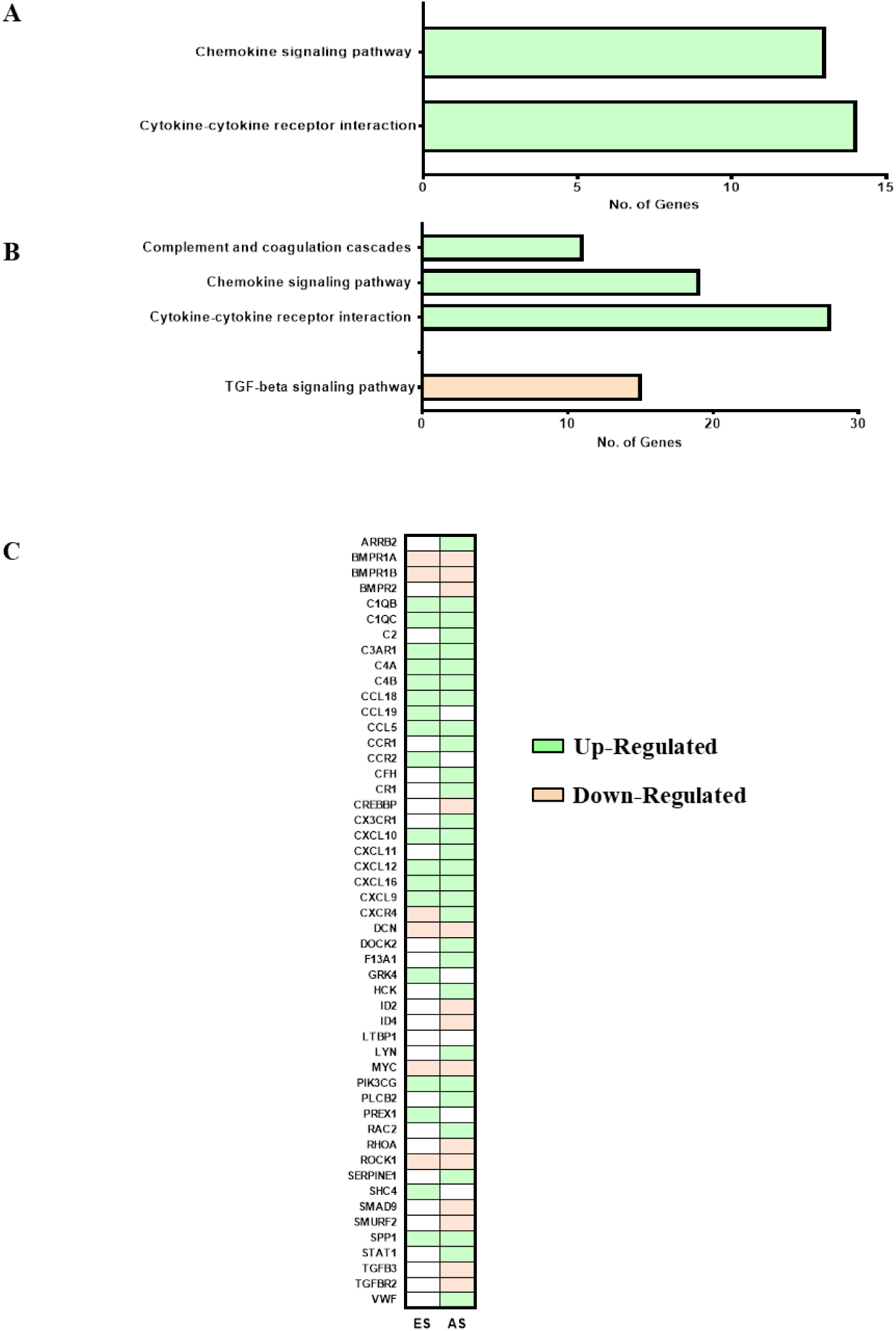
The differentially expressed genes in the plaques are associated with inflammation. Bar graph showing significantly-enriched inflammation associated KEGG pathways of differentially expressed genes in early stage (ES)**(A)** and advanced stage (AS)**(B)** atherosclerotic plaques. The KEGG pathway analysis was done using online database, DAVID. **(C)** List of differentially expressed genes functioning in inflammation pathways mentioned in A and B.*n=2(each with sample pooled from 4 patients); p<0*.*05*

Few of these 19 genes were also found to be up-regulated in AS plaques while the expression ofremaininggenes was found only in ES plaque; for example, PREX1, SHC4, and CCR2**(Figure 3A and 3B)**. These ES plaque specific genes function in regulating the signalling pathways for chemotaxis and migration of various immune cells. Since these genes show plaque specific up-regulation, they can be used as biomarkers to indicate the presence of ES plaque in patients.

**FIGURE 3:**
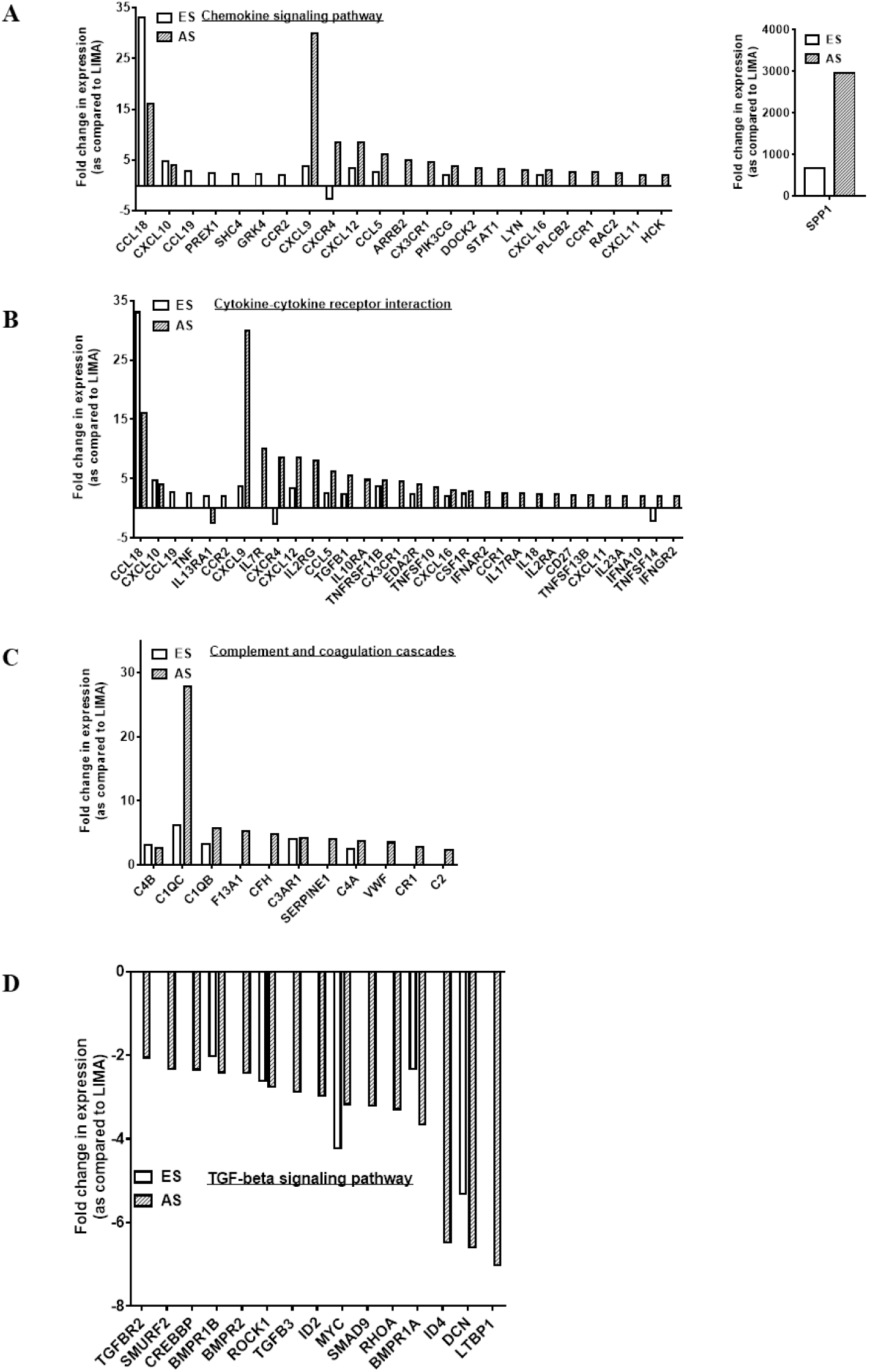
Expression of various inflammatory pathways-associated genes are altered in atherosclerotic plaques. Graphs showing microarray-analysis basedfold change in expression of genes in early stage (ES) and advanced stage (AS) atherosclerotic plaques as compared to (Left Internal Mammary Artery) LIMA. Using online databse DAVID the genes were found to be involved in chemokine signalling pathway **(A)**, Cytokine-cytokine receptor interaction **(B)**, Complement and coagulation cascades **(C)**, and TGF-beta signalling pathway **(D)**.*n=2(each with sample pooled from 4 patients); p<0*.*05*

### AS Plaques are also associated with up-regulation of inflammatory mediators

Upon pathway analysis for up-regulated genes in AS plaques, 48 of these genes were found to be involved in complement and coagulation cascades along with chemokine signalling pathway and cytokine-cytokine receptor interaction **(Figure 2B, 2C, Supplementary Table 2)**. Among the genes functioning in chemokine signalling pathway and cytokine-cytokine receptor interaction, 23 were found to be up-regulated specifically in AS plaque**(Figure 3A and 3B)**. We also observed that the few genes functioning in chemokine signalling pathway and cytokine-cytokine receptor interaction, whose expression was enhanced in both ES and AS plaque, showed higher fold change in the later stage**(Figure 3A and 3B)**. Among these genes are SPP1, CXCL9, CXCL12, and CCL5.

The up-regulated genes involved in Complement and CoagulationCascade include C1QB, C1QC, CFH and C2**(Figure 2B and 3C, Supplementary Table 2)**. These genes function in classical complement pathway which is activated not only during bacterial infections but also in the absence of proper immune regulatory mechanisms. Therefore, in AS plaque, the activation of this system might be involved in the clearance of damaged cells.In summary, the AS plaque is not only associated with a greater number of highly expressed genes but also shows higher fold change of commonly up-regulated genes as compared to ES plaques. Examining the expression levels of such genes in peripheral blood of patients would suggest the prevalence of plaque stage and support the clinicians in further management of the disease.

### AS plaques are associated with down-regulation of anti-inflammatory mediators

The inflammatory response must be actively terminated when no longer needed to prevent unnecessary “bystander” damage to tissues. Failure to do so results in chronic inflammation, and cellular destruction. We found similar observation in AS plaques; 15 down-regulated genes were involved in anti-inflammatory-TGF-beta signalling pathway**(Figure 2D, Supplementary Table 2)**.The loss of expression of some of these genes like BMPRII (receptor of the TGF-beta) and DCN (cellular matrix proteoglycan that binds to TGF-beta) has been reported in human coronary arteries with advanced atherosclerotic lesions further supporting our observations and also suggests an unchecked hyper-inflammatory condition in AS plaques[26]. Few of these genes like BMPR1B, ROCK1, and DCN were also found to be down-regulated in ES plaque as well and their fold change difference was similar to that found in AS plaque**(Figure 3D)**.

### Distinct miRNA profiles in ES and AS plaque

The expression levels of 132 miRNAs differed in the two plaque tissues relative to LIMA. However, the variation in the expression of these miRNAs was insignificant in our data. This might be due to small sample size. Of these miRNAs, 38 were up-regulated and 62 were down-regulated in the ES plaques when compared to LIMA **(Figure 4A and B)**. In AS plaques, 40 miRNAs were up-regulated and 30 miRNAs were down-regulated as compared to LIMA **(Figure 4A and B)**. Also, 5 miRNAs were up-regulated and 29 miRNAs were down-regulated in both the plaques **(Figure 4A and B)**. Recently, the down-regulation of miR-143 has been observed in advanced coronary atherosclerotic plaques[27]. Mir-143 is thought be involved in cardiac morphogenesis and cancer[28]. In human aortic aneurysms the expression of mir-143 has also been found to be significantly decreased when compared to controls[29]. In our study we report down-regulation of this miRNA not only in AS plaque but also in ES plaque. Interestingly the fold change was much lower in ES plaque (−40) than AS plaque (−4), suggesting its possible role in altering vascular smooth muscle cells morphology early during atherosclerosis.

**FIGURE 4:**
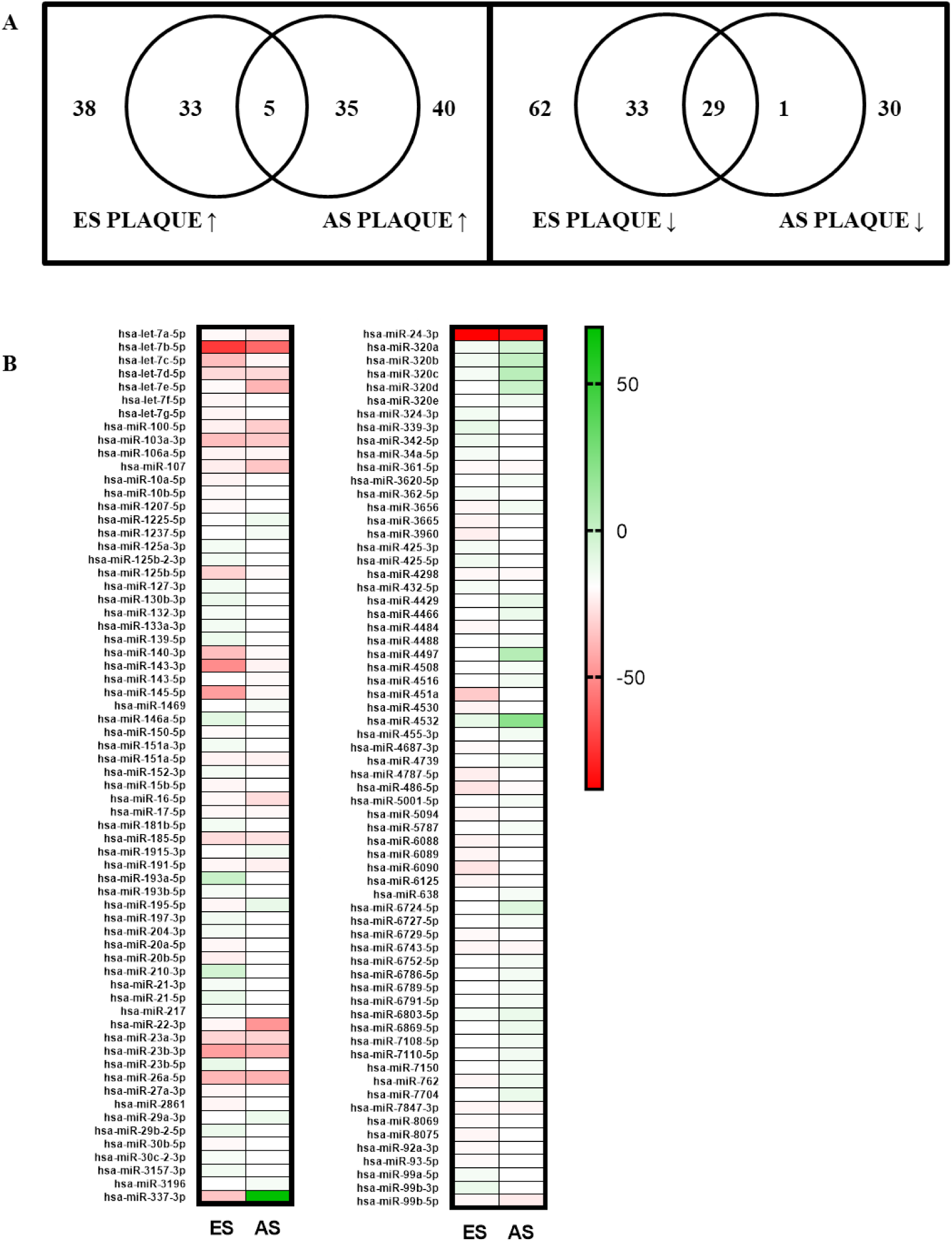
Atherosclerotic plaques are associated with alterations in miRNA expression. **A)** Venn diagram of differentially expressed miRNAsas found in microarray analysis in early (ES) and advanced stage (AS) atherosclerotic plaques as compared to Left Internal Mammary Artery (LIMA). **(B)** Heat-map of miRNAs differentially expressed in the two plaques.*n=2(each with sample pooled from 4 patients)*

The twomiRNAs that were up-regulated in ES showed down-regulation in AS plaque. Of these, miR-22 has been found to be reduced in human arteries from arteriosclerosis obliteran[30]. The down-regulation of miR-22 increases expression of pro-inflammatory cytokines and therefore suggests creation ofunfavourable pro-inflammatory situation that promotes plaque formation[31]. We also found 4 miRNAs, down-regulated in ES plaque, to be up-regulated in AS plaque. In summary, the miRNA expression analysis shows association of atherosclerotic plaques with variable expression of different miRNAs and the presence of similar trend of expression of these miRNAs in ES and As plaques can be used as potential biomarker.

### Negative correlation between miRNAs and mRNAs profile in atherosclerotic plaque

The small non-coding RNAs-miRNAs are known to regulate expression of various genes by inhibiting their translation. In order to understand the role of miRNAs in atherosclerosis associated inflammation, using miRWalk database we first predicted the miRNAs for the inflammatory mediators that were up-regulated in ES and AS plaque as compared to LIMA. Next, these miRNAs were then overlapped with the miRNAs that were found to be down-regulated in the micro-array analysis of respective tissue. Similar methodology was employed for down-regulated inflammatory mediators and the miRNAs targeting them. In this manner, miRNA-target gene interaction pairs of reverse association in the two plaque tissues were obtained. We found 3 such interacting pairs with up-regulated mRNAs and down-regulated miRNAs**(Supplementary Table 3)**. 2 of these pairs included miRNAs (hsa-miR-125b-5p and hsa-miR-23a-3p) down-regulated in AS plaque. Interestingly, the target gene of hsa-miR-125b-5p and hsa-miR-23a-3p-TNFAIP3 was also found to be up-regulated in the AS plaque. The remaining interaction pair included miRNA, hsa-miR-22-3p and its target CSF1R. In both the plaques, this miRNA was down-regulated while its target gene was up-regulated**(Supplementary Table 3)**. The opposite trend of expression of above mentioned miRNAs and mRNAs in patients’ blood can further support in diagnosis of plaque stage.

### Distinct mRNA/miRNA profiles are detected in patients’ blood

With an effort to use the identified mRNAs as biomarkers for detection of atherosclerotic condition, we quantified the levels of few mRNAs in patients’ blood by qRT-PCR and compared with that of healthy blood samples. The panel included the mRNAs that showed altered expression in both the plaques and specifically in ES or AS plaques **(Supplementary Table 4)**. The qRT-PCR result showed up-regulation of genes that were involved in chemokine signalling pathway, cytokine-cytokine receptor interaction and complement and coagulation cascades while the expression of genes that were functioning in TGF-beta signalling pathway was found to be lowered as compared to healthy controls **(Supplementary Figure 4A)**.We also performed QRT-PCR for the miRNA-mRNA pairs found in our study **(Supplementary Table 3)**. Expression pattern similar to the microarray analysis was found.

However, the mRNAs and miRNAs showed non-significant *p* value. Increasing the sample size may improve the significance of our result. These results support the presence of differentially expressed miRNA and mRNA in patients’ blood and their use as potential biomarkers for assessing the stage and risk of atherosclerosis.

## Discussion

Atherosclerosis is a chronic inflammatory condition of arteries. Various effector immune mechanisms and signalling networks are involved in the pathology of atherosclerotic plaque formation[1, 2, 4, 6, 32].Elevated levels of inflammatory cytokines and chemokines have been observed in various studies on affected cardiac tissues[11, 12, 14–17]. However, studies on differential expression of immune/ inflammatory genes in plaques of early versus advancedstages remain scant. Volger et al, 2007, studied gene expression profiles of endothelium from human large arteries having focal atherosclerosis of the ES or AS[17]. The transcriptomes of the lesional arteries were compared with the transcriptomes of their unaffected sides, thus limiting genetic and external confounders. The authors, however, collected samples post-mortem after disease. Post-mortem tissues undergo significant changes in its physical and chemical compositions possibly altering the profile of mRNA stability retrieval and amplification required for transcriptome profiling[33]. Therefore, we performed microarray analysis of ES and AS plaque as well as healthy LIMA tissue obtained surgically from the same live individuals undergoing CABG Surgery.

Here, we attempted to understand the differential expression profiles of immune response related genes in early versus advanced atherosclerotic plaques. We found that the ES and AS plaques are associated with differential expression of various mRNAs and miRNAs, suggesting that the transcriptional regulations and responses in the initial stage of plaque formation may contribute in the subsequent pathophysiology of atherosclerosis **(Supplementary Figure 2)**. In both the plaques, significant numbers of genes were found to be involved in more than one arm of the immune system. Interestingly, few of these genes have already been reported to be involved in atherosclerosis pathology, supporting the validity of our data. Examples of such genes are SPP1, CXCL9, CCL18, FcγRIIIA, etc[20–25]. The higher expression patterns of common genes in AS versus ES plaques suggest that atherosclerosis involves an array of processes where alterations in their profiles shift the balance from ES to AS stage evolution. A prominent example of such common gene is SPP1, required for macrophage chemotaxis, critical in lipid peroxidation, a pathological landmark of atherosclerosis.Our results also unravel the differentially expressedgenes in plaque in stage specific manner, and definitive profiles marking the dominance of early and/or advanced atherosclerotic changes in patients. For example, the chemokine CCL19 was up-regulated only in ES plaque, which might be involved in trafficking of various inflammatory cells at plaque site. On the other hand, AS plaques showed up-regulation of genes involved in stronger inflammation (e.g.-C1QC, C1QB, TYROBP etc).

The over-expression of genes functioning in chemokine signalling pathway in ES plaques suggests initiation of various immune cells recruitment at the affected tissue. For example, CCL18 for naïve T cells, CCL19 for T and B cell migration and CCL5 for blood monocytes, memory T helper cells and eosinophils. The genes (like CCL5, TNF, CXCL10) which are involved in cytokine-cytokine signalling stimulates these incoming immune cells to release various other signalling and pro-inflammatory molecules; for instance, activation of eosinophils by CCL5 and stimulation of monocytes by CXCL10. Therefore, chemotaxis and activation of a diverse immune cell population in ES plaque suggests an onset of inflammation in the affected vascular tissue. The up-regulation of genes coding for different cytokine receptors like IL7R, IL2RG, IFNGR2, and IL17RA only in AS plaque and the absence of expression of their respective cytokines in microarray analysis indicates toward a compensatory phenomenon in which cells attempt to enhance the probability of binding to the scarce cytokines.Also, the suppression of the genes required for TGF-beta signalling in AS plaque and ES plaque suggests a failure of immune-regulatory mechanisms to check the on-going inflammation.

In an attempt to understand the regulation of differentially expressed mRNAs at post-transcriptional level, microarray analysis against miRNAs for both plaques was carried out, followed by identification ofmiRNA-mRNA interaction pairs for the corresponding plaques. Few of the resulting miRNAs like hsa-miR-17, hsa-miR-16, hsa-miR-195, hsa-miR-27a, hsa-miR-20b, etc have already been reported in other studies to be associated with coronary artery disease [34–36]. We found three miRNA-mRNA pairs associated with atherosclerosis. Among them, one pair included hsa-miR-125b and its target TNFAIP3. Hsa-miR-125b act as a negative regulator of inflammatory genes[37]and TNFAIP3 is involved in the cytokine-mediated immune and inflammatory responses. Therefore, the involvement of hsa-miR-125b in inflammation during atherosclerosis can also be proposed via TNFAIP3. The second interaction pair included hsa-miR-22-3p and its target CSF1R. Since CSF1R promotes the release of pro-inflammatory chemokines, the finding that this gene is up-regulated while the miRNA targeting it is down-regulated in ES and AS plaques suggests the role of this miRNA-mRNA pair in atherosclerosis associated inflammation.

Identifying microarray based bio-markers may complement and help in circumventing the invasive techniques for clinical evaluation about the stage of atherosclerotic pathophysiology and its implication in patient prognosis and healthcare. Circulating mRNAs and miRNAs are being investigated as possible biomarkers for early detection and progression of various diseases[38–44]. Moreno et al. (2009) reported that the plasma level of CD163-TWEAK (tumor necrosis factor-like weak inducer of apoptosis) is a potential bio-marker of atherosclerosis[45]. Therefore, differentially expressed mRNAs and miRNAsprofiles identified in the present study may reveal a fingerprint of the presence and/or the dominance of ES and AS plaques. We correlated the same in peripheral blood of the patients with an aim to identify the signature changes in their blood. The pattern of expression difference was similar to that reported in microarray analysis, supporting the hypothesis that the expression levels of common and stage specific mRNA and miRNAs in the blood samples of patients can be used as a biomarker for early detection and monitoring of progression and/or regression of coronary atherosclerosis and thus possibly to evaluate the effectiveness of anti-atherosclerotic therapies on a larger scale. However, we need a proper scoring system for the expression of these stage specific genes so that weightage to ES or AS could be given. Although we cannot know the frequency of the particular plaque stage in the patient, the ratio of the expression of stage specific genes can tell us condition of more prevalent plaque.

## Conclusion

From our results, two distinct mRNA-miRNA pair profiles are evident-

Panel 1-SPP1↑, CCL18↑↑, CXCL9↑, CXCL12↑, IL13AR1↑, TNFSF14↓, CXCR4↓, CCL19↑, TNF↑, C1QC↑, C1QB↑

Panel 2-SPP1↑↑, CCL18↑, CXCL9↑↑, CXCL12↑↑, IL13AR1↓, TNFSF14↑, CXCR4↑, IL7R↑, IL2RG↑, ARRB2↑, IL10RA↑, C1QC↑↑, C1QB↑↑

Former has a high correlation with ES plaque and later with AS plaque. Such profiles are also reflected in the peripheral blood. Therefore, we propose that ratios of these profiles may signature the proportion of advanced pathology in patients. This might supplement the imaging analysis of atherosclerotic plaques and enable clinicians to better assess the disease state and plan for clinical management accordingly.

## Data Availability

Raw data are available from the European Bioinformatics Institute ArrayExpress data repository
(accession number-E-MTAB-10052 (mRNA data); E-MTAB-10051 (miRNA data)).

https://www.ebi.ac.uk/arrayexpress/experiments/E-MTAB-10052/

https://www.ebi.ac.uk/arrayexpress/experiments/E-MTAB-10051/

## Supplementary Tables

**Supplementary Table 1:**
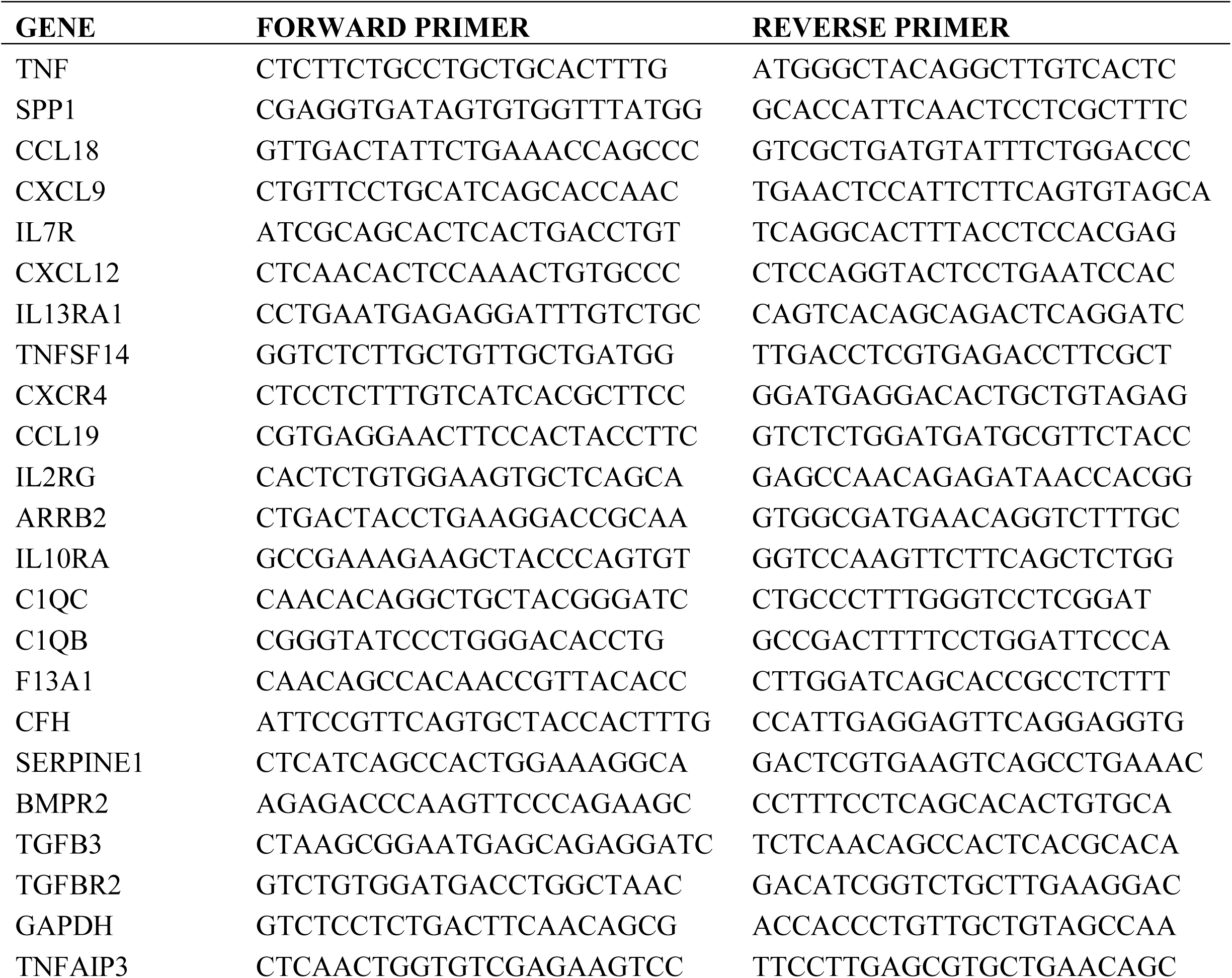
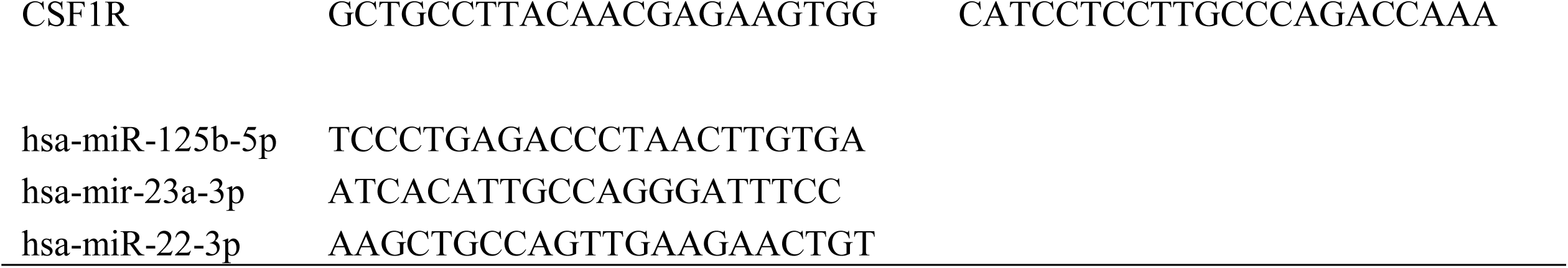
The list of primers used for qRT-PCR.

**Supplementary Table 2:**
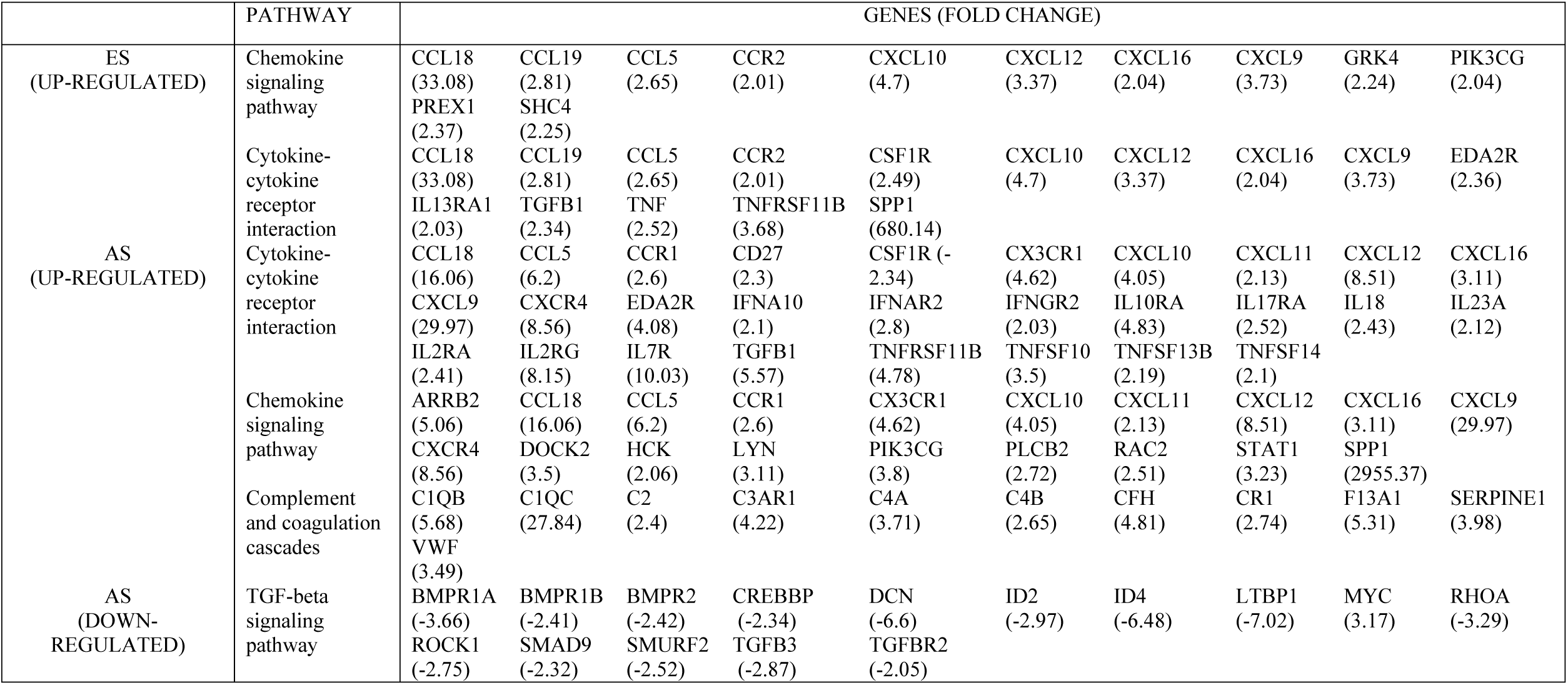
Significantly-enriched inflammation associated KEGG pathways of differentially expressed genes in early stageand advanced stage atherosclerotic plaque.

**Supplementary Table 3:** miRNA-target gene interaction pairs of reverse association in the two plaque tissues.

|  | miRNA | Fold Change<br>vs LIMA | Target<br>mRNA | Fold Change<br>vs LIMA |
| --- | --- | --- | --- | --- |
| ADVANCED | hsa-miR-125b-5p | -2.26 | TNFAIP3 | 4.88 |
| STAGE | hsa-miR-23a-3p | -15.88 | TNFAIP3 | 4.88 |
| EARLY STAGE | hsa-miR-22-3p | -3.32 | CSF1R | 2.49 |
| ADVANCED |  |  |  |  |
| STAGE | hsa-miR-22-3p | -36.7 | CSF1R | 2.95 |

**Supplementary Table 4:** The panel of genes proposed in the study to be used as biomarker for diagnosis of plaque stage.

|  | ES | AS |
| --- | --- | --- |
| Chemokine signalling pathway and<br>Cytokine-cytokine receptor interaction |  |  |
| SPP1 | ↑<sub>a</sub> | ↑↑ |
| CCL18 | ↑↑ | ↑ |
| CXCL9 | ↑ | ↑↑ |
| CXCL12 | ↑ | ↑↑ |
| IL13RA1 | ↑ | ↓<sub>b</sub> |
| TNFSF14 | ↓ | ↑ |
| CXCR4 | ↓ | ↑ |
| CCL19 | ↑ |  |
| TNF | ↑ |  |
| IL7R |  | ↑ |
| IL2RG |  | ↑ |
| ARRB2 |  | ↑ |
| IL10RA |  | ↑ |

| Complement and coagulation cascades |  |  |
| --- | --- | --- |
| C1QC | ↑ | ↑↑ |
| C1QB | ↑ | ↑↑ |
| F13A1 |  | ↑ |
| CFH |  | ↑ |
| SERPINE1 |  | ↑ |
| TGF-beta signaling pathway |  |  |
| BMPR2 |  | ↓ |
| TGFB3 |  | ↓ |
| TGFBR2 |  | ↓ |

*<sup>a</sup>up-regulated, <sup>b</sup>down-regulated*

## Supplementary Figures

**Supplementary Figure 1:**
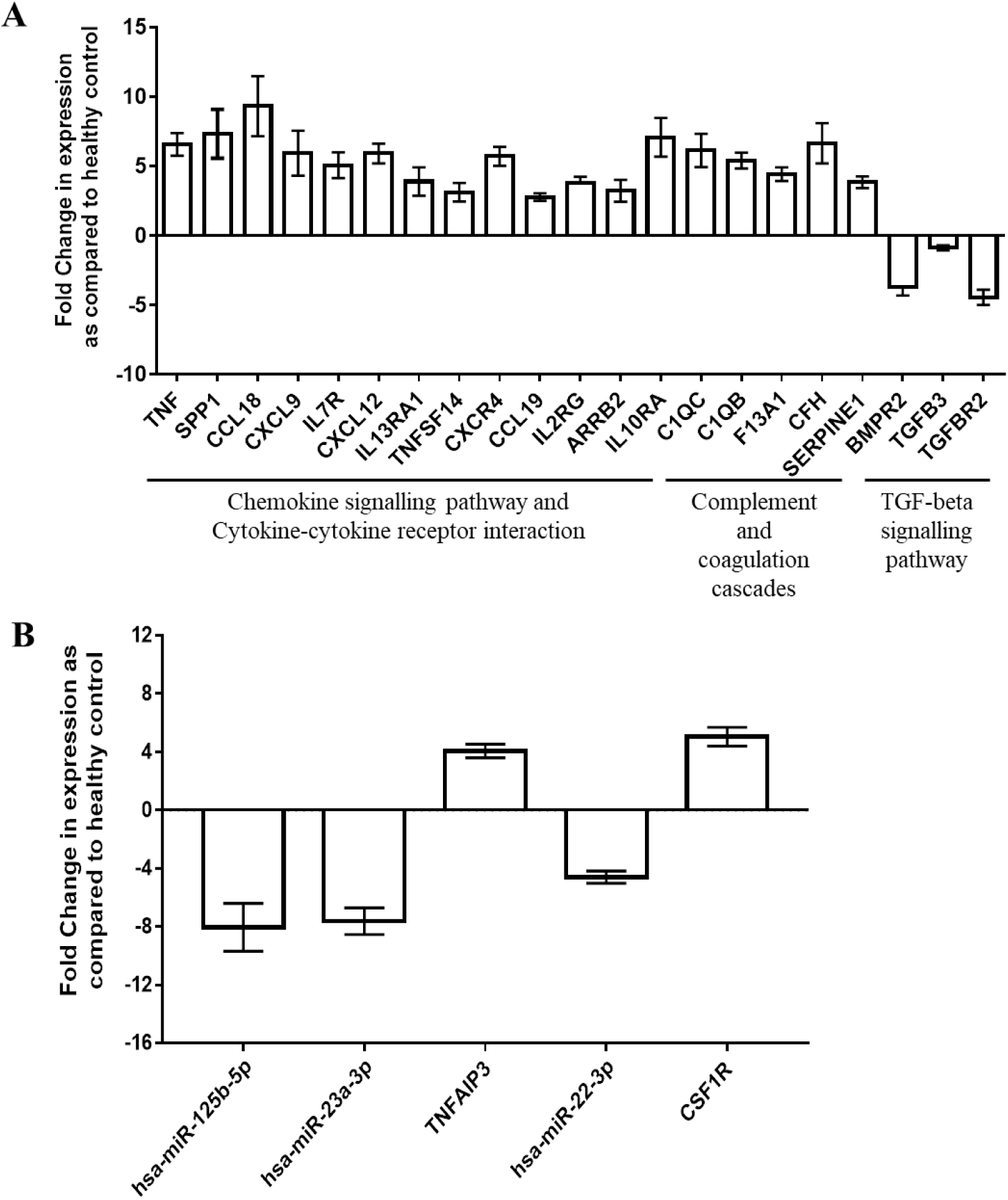
The Expression of mRNA/miRNA is also altered in peripheral blood of patients. **(A)** QRT-PCR based quantification of mRNAs of proposed biomarker panel in peripheral blood of patients and comparison with healthy control. **(B)** QRT-PCR based quantification of identified miRNA-mRNA interaction pairs in peripheral blood of patients and comparison with healthy control. GAPDH mRNA was used for normalization. *n=13 (8 patients; 5 healthy control); Error bar indicates standard error mean. Student’s t test was used to determine statistical significance. p= non-significant*

**Supplementary Figure 2:**
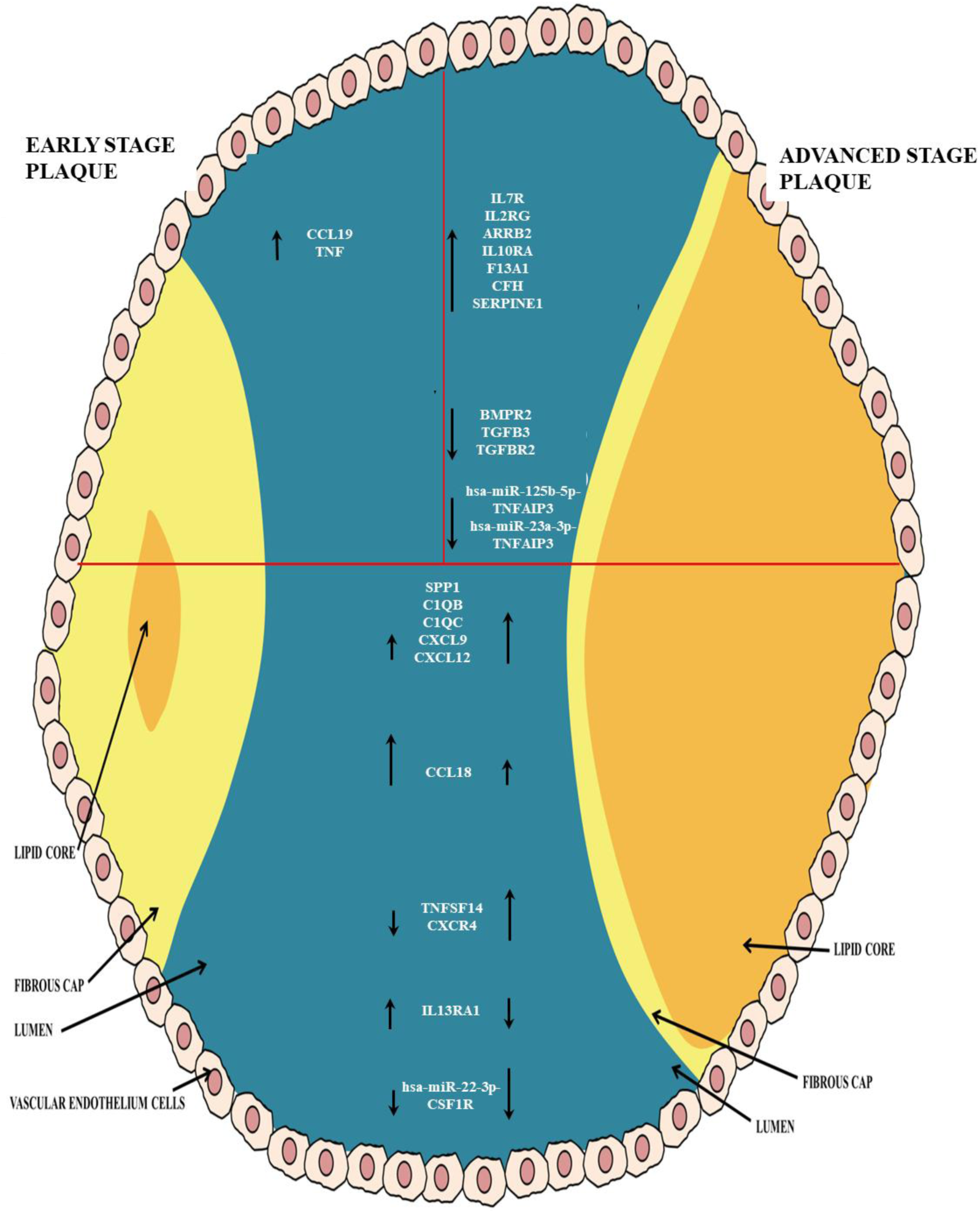
Summary of the microarray analysis of mRNA and miRNA expression pattern in early and advanced stage atherosclerotic plaques as compared to LIMA tissue. The upper left panel represents the differentially expressed in early stage plaques only. The upper right panel represents the differentially expressed mRNA and miRNA along with miRNA-mRNA pairs in advanced stage plaques only. The lower panel shows mRNA, miRNA and miRNA-mRNA pairs differentially expressed in both the plaques.

## Notes

**Conflict of interest:** The authors declare that they do not have anything to disclose regarding conflict of interest with respect to this manuscript.

### Competing Interest Statement

The authors have declared no competing interest.

### Funding Statement

no external funding

### Author Declarations

Institutional Ethics Committee All India Institute of Medical Sciences New Delhi

